# Exploring the potential of a saliva-based, RNA-extraction-free PCR test for the multiplexed detection of key respiratory pathogens

**DOI:** 10.1101/2023.10.04.23296240

**Authors:** Orchid M. Allicock, Tzu-Yi Lin, Katherine T. Fajardo, Devyn Yolda-Carr, Maikel S. Hislop, Jianhui Wang, Denora Zuniga, William Platt, Beth Tuohy, Anne L. Wyllie

**Author notes:** **Corresponding author: Anne Wyllie**, Yale School of Public Health, LEPH 823, 60 College St, New Haven, CT 06510.

## Abstract

**Introduction:** Efforts to diagnose and monitor transmissible respiratory infections can be impaired by invasive or resource-intensive sample collection. Having extensively demonstrated the feasibility of saliva for SARS-CoV-2 detection, we sought to validate its potential for other common upper respiratory tract pathogens.

**Methods:** We modified our RNA-extraction-free SARS-CoV-2 PCR test for multiplexed detection of influenza A/B (IAV/IBV), respiratory syncytial virus (RSV) and human metapneumovirus (hMPV). Stability of virus detection in saliva from virus-positive patients was tested after storage at +4°C, room temperature (∼19°C), 30°C and 40°C for up to 7 days and through simulated shipping conditions. De-identified saliva samples were collected from individuals (≥18 years) with respiratory symptoms who were undergoing nasal-swab-based testing for SARS-CoV-2 (New Haven, CT). Saliva samples from SARS-CoV-2-negative individuals were tested with the multiplexed assay, with and without RNA extraction.

**Results:** The limit of assay detection ranged from 3-6 copies/μl, virus target depending. Detection remained stable after prolonged sample storage at elevated temperatures and through shipping conditions. From the symptomatic testing sites, 1,095 clinical specimens tested SARS-CoV-2-negative. Upon multiplexed testing of their paired saliva, 41 (3.7%) tested positive (IAV, n=20; RSV, n=5; hMPV, n=7). Additionally, upon screening samples in singleplex for pneumococcus, 29 (3%) samples tested positive.

**Conclusion:** Our findings emphasize the adaptability of a low-cost, open-source saliva-based PCR test for common respiratory pathogens, beyond SARS-CoV-2. We demonstrated its utility in symptomatic individuals, identifying viral infection missed when testing focused solely on a singular target, such as SARS-CoV-2.

## INTRODUCTION

Acute respiratory infections (ARIs) are a major health problem globally, and one of the major causes of morbidity and mortality (Vos et al., 2020). They can affect both the upper and lower respiratory tract; severe infections can result in hospitalization. The etiology of ARIs can be either viral or bacterial, with the most common offenders including adenovirus, *Bordetella pertussis*, human bocavirus, coronaviruses, human metapneumovirus (hMPV), human rhinovirus, influenza virus (IAV/IBV), human parainfluenza virus, respiratory syncytial virus (RSV), *Streptococcus pneumoniae* and *Streptococcus pyogenes*, among others (Brandt et al., 1973; Cooney et al., 1975; Edwards et al., 2013; Marom et al., 2014; Miller et al., 2007; Monto, 2002). While vaccines or therapeutics are available for some of these key pathogens (CDC, 2023), as the symptoms for most of their resulting infections are similar, extensive testing is required to identify the etiologic agent before treatment can be recommended, especially in more severe cases.

For the majority of ARIs, antigen and/or PCR-based diagnostics are available, and usually performed with nasopharyngeal swab specimens (Ginocchio & McAdam, 2011). Since 2020, individuals with respiratory symptoms have been encouraged to seek testing as part of COVID-19 control measures, however testing sites and many clinics have been focused solely on testing for SARS-CoV-2 (Hills et al., 2020). Meanwhile, the annual seasonality of IAV/IBV, RSV, and hMPV was disrupted during the COVID-19 pandemic. Cases were virtually absent in the early-to-mid pandemic period of 2020 and 2021 (Brueggemann et al., 2021; Danino et al., 2022; Yuan et al., 2022). As cases reappeared in the later pandemic period, with more severe outbreaks in 2022 and not limited to the usually seasonal periods for each (Kuitunen et al., 2022; Li et al., 2022), with this slowly returning to normal at different rates around the world (Li et al., 2019; Moriyama et al., 2020).

Having demonstrated the potential of saliva to overcome numerous challenges faced during the pandemic for the detection of SARS-CoV-2 (Ott et al., 2021; Vogels et al., 2021) and recognizing its potential for the low-resource discrimination between pathogens(Tan et al., 2022) which produce similar symptom profiles (Laxton et al., 2023), we modified our saliva-based RNA-extraction-free PCR test for SARS-CoV-2 for multiplexed detection of four key respiratory viruses: IAV/IBV, RSV, and hMPV and evaluated its potential application.

## METHODS

### Ethics

Receipt of de-identified, remnant clinical saliva samples was approved by the Institutional Review Board of the Yale Human Research Protection Program (HIC #2000029551). Collection of saliva samples from symptomatic individuals presenting to the Yale Health COVID-19 Clinic was approved by the Institutional Review Board of the Yale Human Research Protection Program (HIC #2000031600). Individuals were informed in writing about the purpose and procedure of the study and consented to study participation through the act of providing a saliva sample.

### Extraction-free detection of respiratory pathogens

A total of 6.5 μL (20 mg/mL) of Proteinase K (New England Biolabs) was added to 50 μL aliquots of saliva in 8-strip tubes. The tubes were placed in a rack and vortexed for 1 minute at 3200 RPM. Samples were heated for 5 minutes at 95°C on Applied Biosystems MiniAmp Thermal Cycler (ThermoFisher Scientific), and then 5 μL of processed saliva (lysate) was used as input for the multiplex RT-qPCR assay.

### Multiplexed PCR assays for the detection of respiratory viruses

We expanded the singleplex SARS-CoV-2 ‘SalivaDirect’ PCR test (Vogels et al., 2021) for multiplexed detection of SARS-CoV-2, IAV/IVB and RSV, using previously published primers and probes (**Table 1**). For this assay, both IAV and IBV probes were labeled with HEX fluorophore, so a positive result would require reflex singleplex testing for virus confirmation. For testing samples of known SARS-CoV-2-negative status, we further modified the assay by substituting the primers and probe for SARS-CoV-2 detection with those targeting hMPV. In both assays, human RNase P (RP) was included for sample quality control.

**Table 1.**
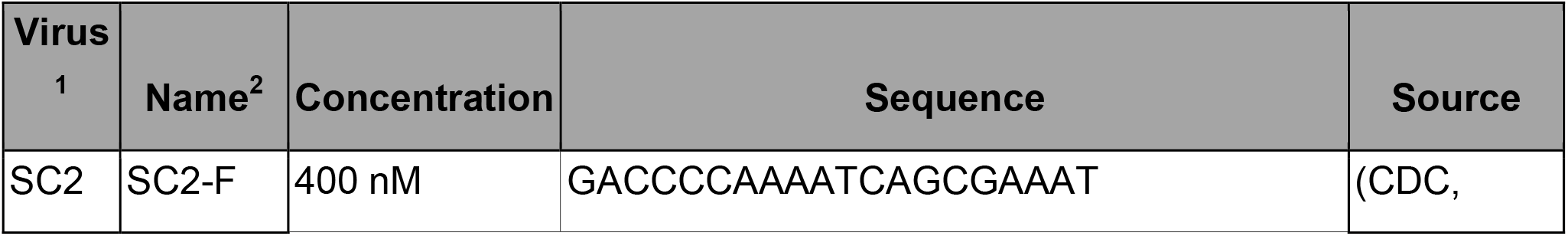
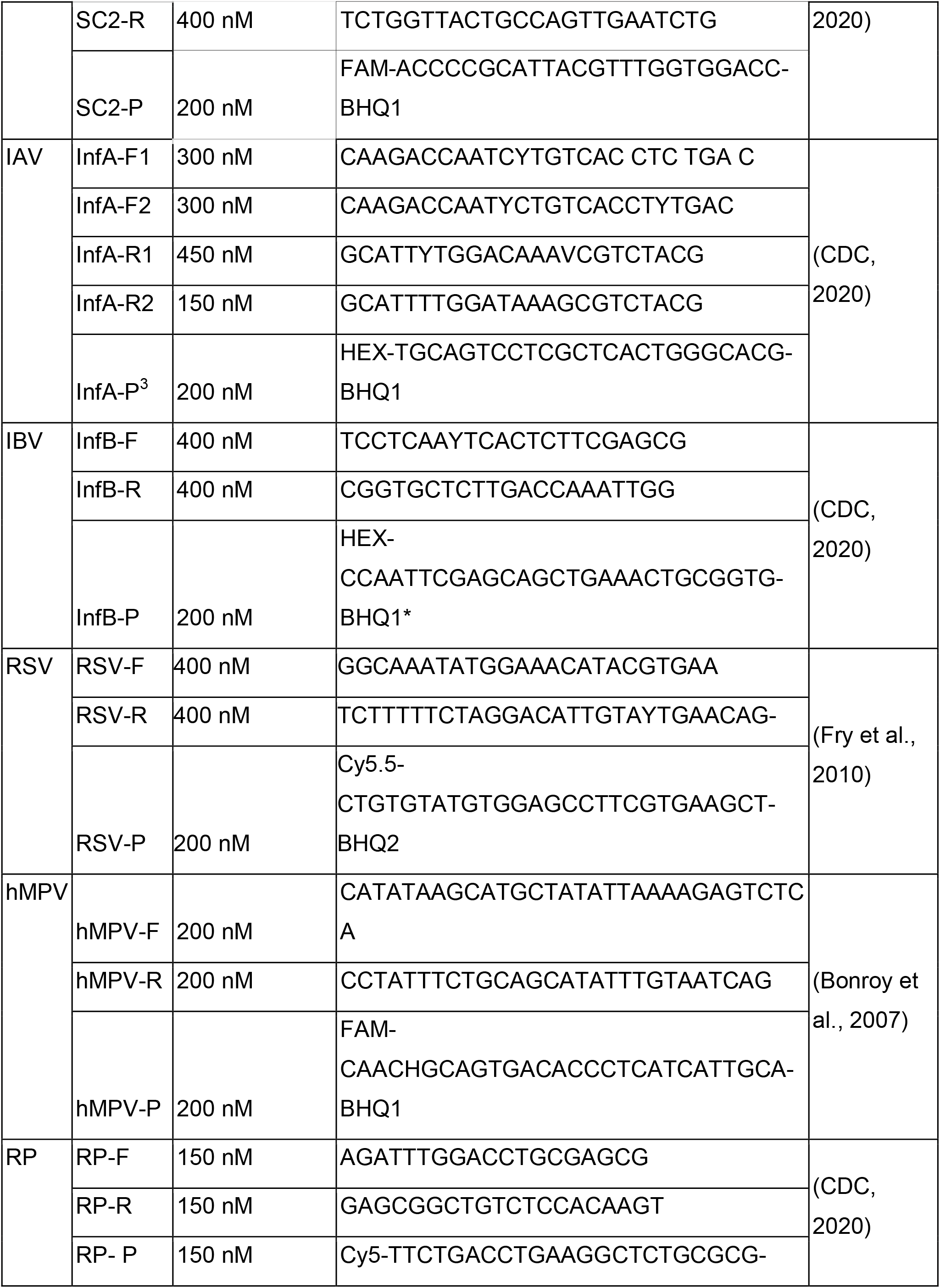
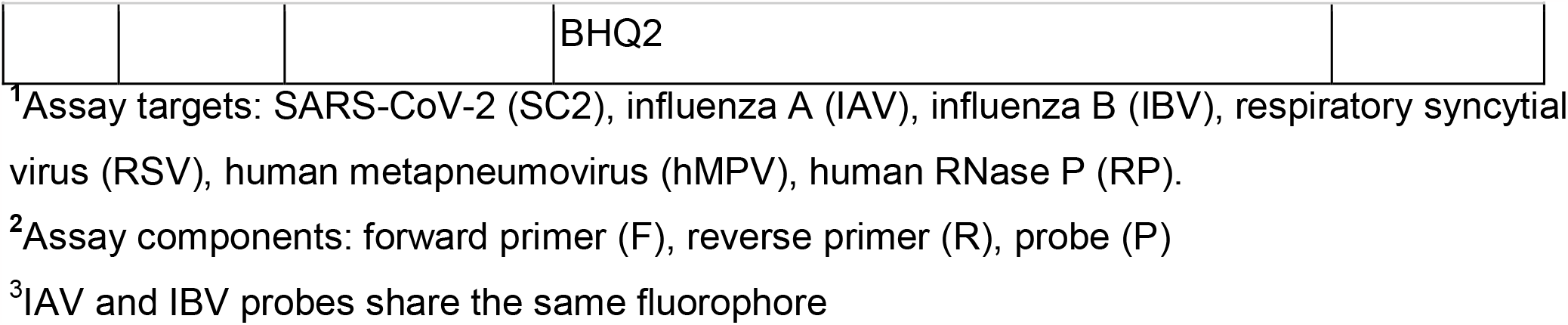
Primers and probes used for the multiplexed qPCR detection of SARS-CoV-2, IAV, IBV, RSV, hMPV and human RNase P (for sample quality control).

### qPCR cycling conditions for the multiplexed detection of respiratory viruses

For each qPCR reaction, 5 μL of template (e.g. extracted nucleic acid or saliva lysate) was tested with Luna® Universal One-Step RT-qPCR Kit (New England Biolabs) using the CFX 96 Touch (Bio-Rad) following the SalivaDirect RT-qPCR protocol (Vogels et al., 2021). The thermocycler conditions consisted of 10 minutes at 52°C, 2 minutes at 95°C, and 45 cycles of 10 seconds at 95°C and 30 seconds at 55°C. Specimens were considered virus-positive if virus-specific Ct <40 and RP Ct <35.

### Assay limit of detection

To determine the preliminary limit of detection (LOD) for each assay, we created a 2-fold dilution series which we tested in triplicate. For IAV, IBV, RSV and RP we used the Exact Diagnostics SARS-CoV-2, Flu, RSV Positive Run Control (catalog #COVFLU; Bio-Rad, Fort Worth, USA). For hMPV we used a synthetic RNA control (#VR-3250SD, ATCC).

### Nucleic acid extraction protocol

Nucleic acid was extracted from 200 μL of saliva and eluted into a final volume of 50 μL using the MagMAX Viral/Pathogen Nucleic Acid Isolation Kit (ThermoFisher Scientific) on the KingFisher Apex (ThermoFisher Scientific), following a modified protocol (Wyllie et al., 2020).

### Stability of virus detection in raw, unsupplemented saliva stored for prolonged periods and at elevated temperatures

We received remnant IAV- and RSV-positive saliva samples from clinical laboratories within the U.S. SalivaDirect laboratory network to test the stability of detection of IAV and RSV in raw (unsupplemented) saliva. This stability was determined by placing aliquots of virus-positive clinical saliva for up to 7 days at 4°C, room temperature (RT, ∼19°C), and 30°C and for up to 3 days at 40°C. Using saliva samples from the New Haven symptomatic testing sites that tested positive for hMPV, we tested the stability of hMPV detection, placing aliquots of saliva for up to 3 days at 4°C, RT, and 30°C. Results were compared to results when samples were tested prior to incubation.

We also evaluated the stability of IAV and RSV detection following conditions replicating those encountered during the shipment of saliva samples from site of collection to the clinical diagnostic laboratory. As recommended by the U.S. Food and Drug Administration (FDA), virus-positive samples were cycled through temperatures chosen to represent possible shipping conditions in summer and winter (ISTA, 2007). For the summer profile, samples were incubated at 40°C for 8 hours, 19°C for 4 hours, 40°C for 2 hours, 30°C for 36 hours, then 40°C for 6 hours. For the winter profile, samples were incubated for 8 hours at -20°C, room temperature (∼19°C) for 4 hours, -20°C for 2 hours, 4°C for 36 hours, then -20°C for 6 hours. Following incubation, saliva specimens were tested as described for the extraction-free protocol, with results compared to when samples were tested prior to incubation.

### Collection and testing of clinical specimens

During the first sampling period (May-July 2022), clinical nasal swab and saliva specimens were collected from the Yale Health COVID-19 testing site (New Haven, CT, USA) from symptomatic individuals. Nasal swabs were self-collected according to standard diagnostic practice. Saliva samples were self-collected as previously described (Allicock et al., 2022). Nasal swab specimens were tested for SARS-CoV-2 at the Yale Pathology Labs (New Haven, CT, USA). During the second sampling period in October 2022-February 2023, remnant clinical saliva specimens from several sites were also tested for SARS-CoV-2 at Yale Pathology Labs. Both sets of saliva samples from individuals testing negative for SARS-CoV-2 were transferred to the research laboratory at the Yale School of Public Health. All specimens were aliquoted and stored at -80ºC until further analysis.

### Singleplex PCR detection of Streptococcus pneumoniae in saliva samples from symptomatic, SARS-CoV-2-negative individuals

All SARS-CoV-2-negative saliva samples from the symptomatic testing site were also screened for the presence of pneumococcus, using the extraction-free approach as described previously and targeting pneumococcal-specific gene, *piaB* (Trzciński et al., 2013; Wyllie et al., 2016).

### Statistical analysis

We used the Bio-Rad CFX Maestro 1.1 V4.1.2435.1219 to analyze and export Ct values. GraphPad Prism 10.0.1 was used for all statistical analyses. The Wilcoxon matched pairs test was used to test for statistical differences between paired samples or conditions. If a virus target was not detected, the Ct value was set to 45. In all statistical tests, *p* ≤ 0.05 was considered significant.

## RESULTS

### Assay limit of detection

The limit of detection (LOD) for each of the virus targets in the multiplexed PCR assay were determined to be 6 virus copies/μl for SARS-CoV-2, IAV/IBV, and RSV. When the primer and probe sequences for SARS-CoV-2 were replaced for those targeting hMPV, the LOD for IAV/IBV, RSV and RP remained unchanged and the LOD for hMPV was determined to be 3 copies/μL.

### Screening of clinical saliva samples

Of the paired nasal and saliva specimens obtained from symptomatic individuals at the Yale Health COVID-19 Clinic, 804 nasal swabs tested negative in the clinical lab (Yale Pathology Labs) for SARS-CoV-2. Using the multiplex PCR assay targeting IAV/IAB, RSV and hMPV, we screened the matching saliva samples that we received from the clinical lab. In total, 17 (2.1%) of 804 saliva samples tested positive for one of the viruses targeted. Of these, the majority of the virus detected was IAV (7/15, 47%), then hMPV (6/17, 35%), then RSV (3/17, 18%). From October 2022 to February 2023, 291 remnant saliva samples from clinical, SARS-CoV-2 testing at Yale Pathology Labs were available for analysis. Of these, 15 (5%) tested positive for one of the targeted viral pathogens. As for the previous period, the majority of the viruses detected were identified as IAV (13/15, 87%), with one positive each of RSV and hMPV (1/15, 6.7% each).

### Stability of IAV and RSV detection in raw, unsupplemented saliva

Using remnant clinical saliva samples from virus-positive individuals, we observed stable detection of IAV and RSV for up to 3 days when stored at 4°C (IAV: ΔCt = 0.79, *p* = 0.31; RSV: ΔCt = 1.59, *p* = 0.063), room temperature (IAV: ΔCt = 0.79, *p* = 0.31; RSV: ΔCt = 0.14, *p* = 0.63) and 30°C (IAV: ΔCt = 0.75, *p* = 0.25; RSV: ΔCt = 0.28, *p* = 0.88) without the addition of preservatives (**Figure 1**). Detection of IAV also remained stable for up to 3 days at 40°C (ΔCt = 0.95, *p* = 0.13), but we observed a decline in the detection of RSV during this period (ΔCt = 6.05, *p* = 0.03). Additionally, we observed stable detection of IAV and RSV following incubation for 56 hours through cycling conditions designed to represent potential temperatures encountered when shipping samples through the post in both winter (IAV: ΔCt = 0.34, *p* = 0.87; RSV: ΔCt = 0.10, *p* = 0.94) and summer (IAV: ΔCt = 0.66, *p* = 0.49; RSV: ΔCt = 0.45, *p* = 0.34).

**Figure 1.**
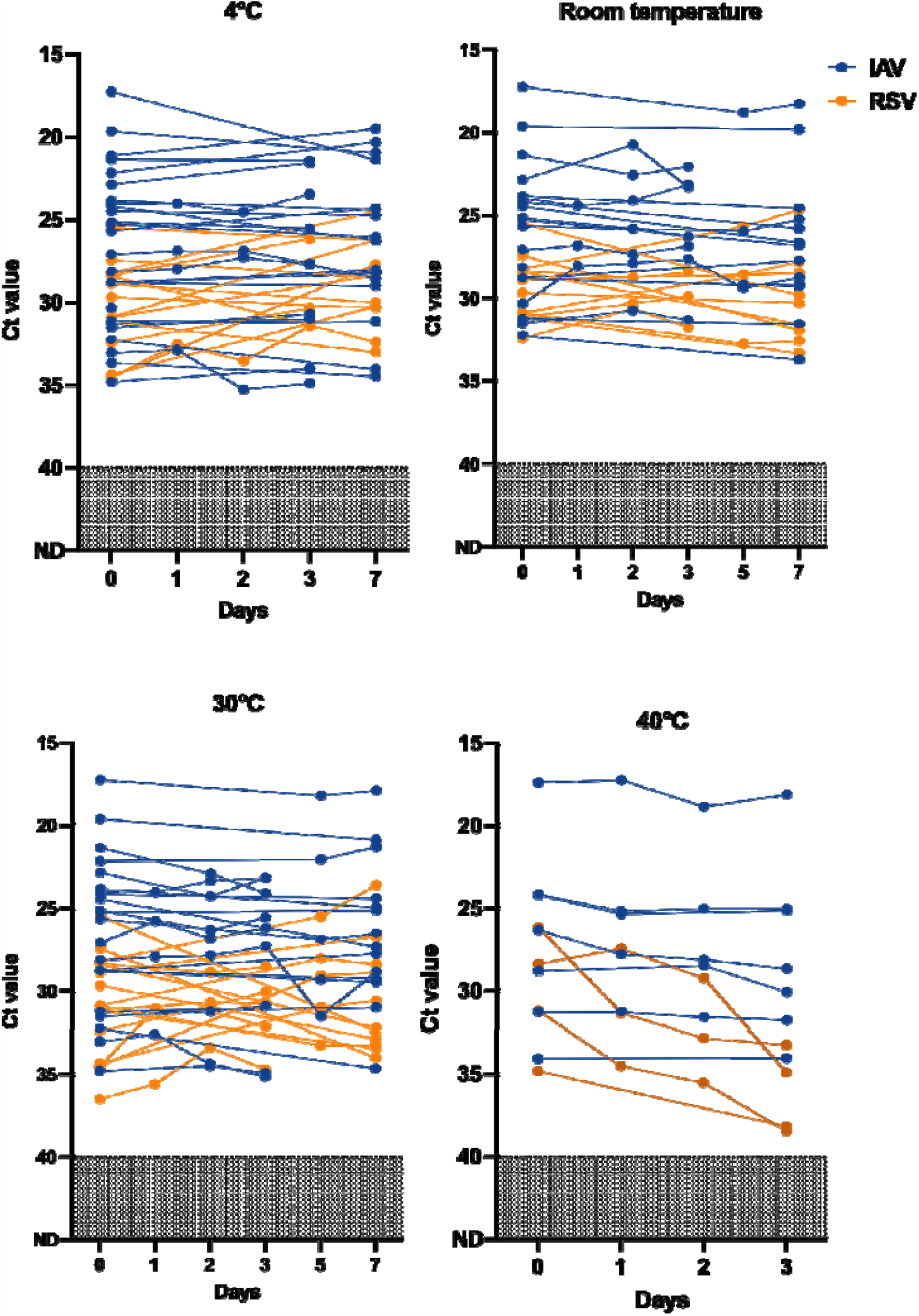
Detection of influenza A virus (IAV) and respiratory syncytial virus (RSV) in raw, unsupplemented saliva stored at 4ºC, room temperature (∼19ºC), 30ºC and 40ºC for up to 7 days. Remnant, de-identified saliva samples were stored without preservative for up to 7 days. Detection of both IAV and RSV remained relatively stable at 4°C, room temperature and 30°C. Detection of IAV also remained stable for up to 3 days at 40°C, but detection of RSV was less stable. Each dot joined by a solid line represents the same sample and the days it was tested on. Samples that remained not detected (ND) by qPCR after 45 cycles are depicted as Ct = 45. See Table S3 for data.

### Stability of hMPV detection in raw, unsupplemented saliva

With the hMPV-positive saliva samples that we identified from the symptomatic testing site, we explored the stability of virus detection in raw, unsupplemented saliva. We confirmed stable detection of hMPV when stored at 4°C (*p* > 0.99), room temperature (*p* = 0.17) and 30°C (*p* > 0.99) without the addition of preservatives (**Figure 2**).

**Figure 2.**
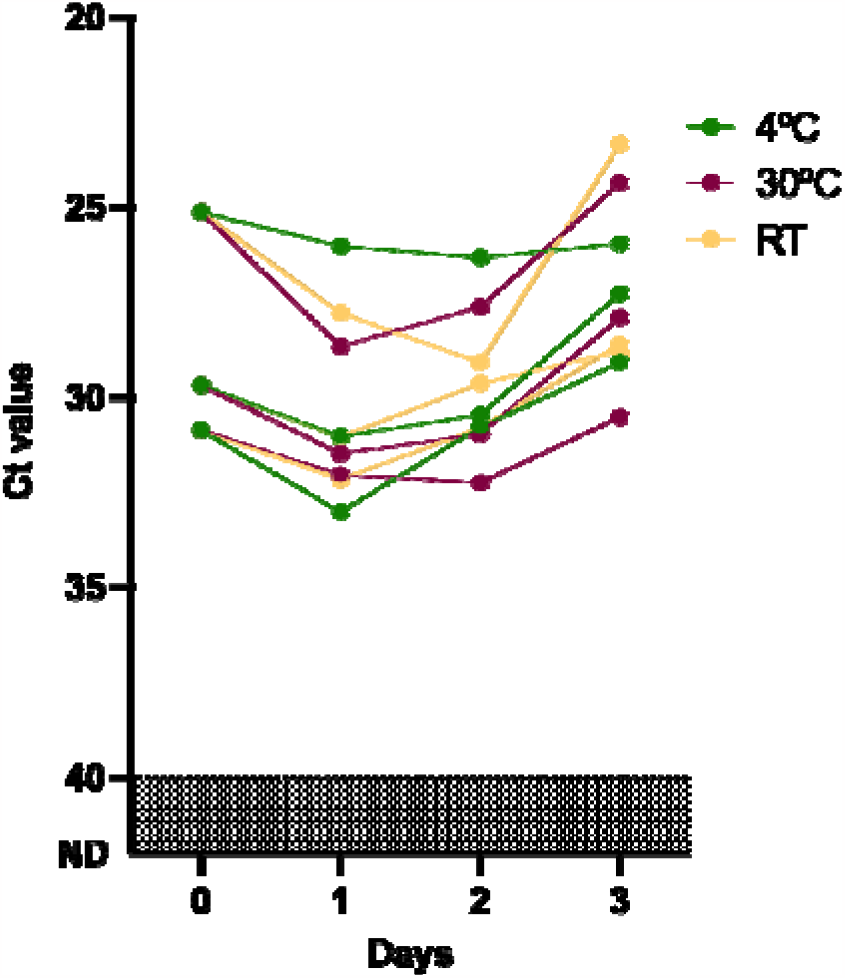
Stable detection of hMPV in raw, unsupplemented saliva stored at 4°C, room temperature (∼19°C) and 30°C and for up to 3 days. Saliva samples from symptomatic SARS-CoV-2 negative individuals, which tested positive in our multiplexed PCR for hMPV, were stored for up to 3 days and re-tested daily. We confirmed stable detection of hMPV when stored at 4°C, room temperature and 30°C without the addition of preservatives. Each dot joined by a solid line represents the same sample and the days it was tested on. See Table S3 for data.

### Sensitivity of RNA-extraction-free sample processing compared to nucleic acid extraction

We compared the Ct values obtained for IAV, RSV and hMPV from the virus-positive clinical saliva samples when using the extraction-free and standard nucleic acid extraction protocols (**Figure 3**). We observed no significant difference between the Ct values obtained from samples positive for IAV (ΔCt = 1.69, *p* = 0.13) or RSV (ΔCt = 2.77, *p* = 0.75), with 100% positive agreement between the different extraction methods. However, for the detection of hMPV, we found TNA extracted from the saliva samples to be significantly more sensitive as compared to testing of the saliva lysates (ΔCt = 7.2; *p* = 0.03), with only 83% agreement between the two methods.

**Figure 3.**
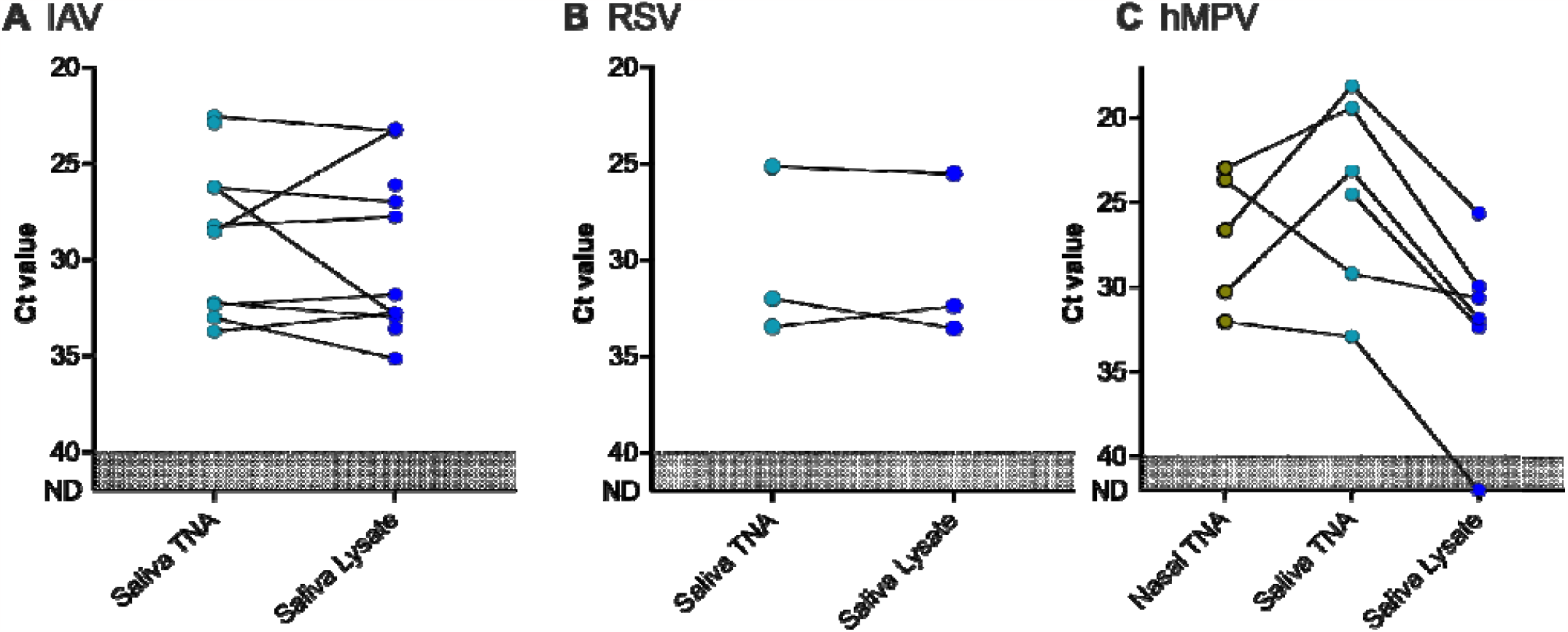
Comparison of the sample processing methods standard nucleic acid extraction (TNA) and the extraction-free (lysate) protocols for the detection of IAV, RSV, and hMPV in saliva. TNA and lysates for each sample were tested using the multiplex PCR assay targeting IAV, IBV, RSV and hMPV. There was no significant difference between the different processing methods for IAV and RSV. For hMPV, we also compared TNA derived from nasal swabs (nasal TNA), and there was a significant difference between the saliva TNA and lysate. Dots connected by a solid line represent an individual sample (see table S2 for data). Samples that remained not detected (ND) by qPCR are depicted as Ct = 45.

Additionally, for the 6 saliva samples that were identified as hPMV-positive, we were able to obtain 5 of their paired, remnant nasal swab samples from the clinical lab. Aliquots of the nasal samples were also extracted, and we compared hMPV detection among the saliva lysates, the TNA extracted from the saliva samples and the nasal swabs. There was no significant difference between the nasal TNA and saliva lysates (*p* = 0.44).

### Application of the extraction-free approach for the detection of Streptococcus pneumoniae in SARS-CoV-2-negative individuals

When the 804 saliva samples from symptomatic individuals, collected during the first sampling period were screened for the pneumococcal gene *piaB*, we identified 29 (3.6%) positive samples. Three of the positive samples had also tested virus-positive (one each of IAV, RSV and hMPV). We detected more pneumococcus-positive samples during the second sampling period, which wasn’t restricted to symptomatic individuals (12/291, 4.1%), however three co-infections with IAV were detected.

## DISCUSSION

The concept of using saliva as a diagnostic tool for respiratory pathogens has gained widespread attention since being demonstrated as an acceptable and sensitive sample type for the detection of SARS-CoV-2 during the COVID-19 pandemic (Tobik et al., 2022). Adding to the global success of many saliva-based SARS-CoV-2 tests, a recent study by Ramirez et al. (Ramirez et al., 2023) demonstrated the utility of saliva for RSV diagnosis. They observed that its inclusion alongside the gold-standard NPS resulted in a twofold increase in the diagnostic yield when compared to NPS alone. This finding perhaps should have been expected, based on the wealth of data that existed prior to the COVID-19 pandemic suggesting the potential of saliva for the sensitive detection of numerous other respiratory pathogens (Laxton et al., 2023). Based on this, and the shifting attitudes towards saliva, we explored the expansion of our open-source, saliva-based RNA-extraction-free PCR test for SARS-CoV-2 for its ability to detect other common respiratory pathogens, namely, influenza virus, RSV and hMPV.

Our initial validation of this approach highlights the potential for saliva to become a valuable tool for community surveillance and testing. In the development of the multiplexed assay, we were able to easily incorporate qPCR assays previously published and used by others. After establishing an assay LOD comparable to many of the diagnostic assays which were authorized for the detection of SARS-CoV-2, we tested its potential in real world applications, screening saliva samples collected from 1,095 SARS-CoV-2-negative individuals displaying symptoms of respiratory infections. With this, we unveiled viral infection in 32 individuals which were otherwise unreported when the symptomatic testing program focused solely on SARS-CoV-2.

Typically, the respiratory virus season occurs from mid-late fall to early spring in temperate locations. The first sampling period of this study fell from late spring to summer, and the second sampling period began late fall and lasted until winter. In line with this, our detection rate for IAV went from 0.8% during the first period, to 4.5% during the second period, which was higher than the influenza regional baseline of 2% symptomatic cases (CDC, 2022). The detection rate for RSV however, was well below the 3% threshold (Hamid et al., 2023) for both sampling periods, but case rates are typically higher in children (McLaughlin et al., 2022), who were not sampled in this study. As influenza and RSV return to their regular seasonal patterns, and SARS-CoV-2 outbreaks continue, the need for multiplexed testing of symptomatic individuals becomes increasingly crucial. This streamlined testing approach provides a cost-effective approach to distinguish the causative agent responsible for the similar symptom profiles that these viruses produce. The existing respiratory panels are resource-intensive, costly, and usually reserved for hospital settings. Simplified, multiplexed PCR testing offers a cheaper alternative, saving hospitals and clinics important resources.

Critical to the development of this low-cost, saliva-based test, we confirmed that the detection of the targeted viruses remained stable in raw, unsupplemented saliva for up to 7 days at 30°C, similar to what we previously observed for SARS-CoV-2 (Ott et al., 2021). While both IAV and RSV detection remained stable throughout simulated summer and winter shipping conditions, we were also surprised to find stable detection of IAV following 3 days at 40°C. Although the detection of RSV was less stable at 40°C, it suggests that short periods of cold-chain-free transport may be permissible, easing the burden on low-resource settings. The ability to not only detect IAV, RSV and hMPV in saliva, but also after different storage and shipping conditions, without more expensive collection devices containing proprietary reagents, has important implications for surveillance initiatives especially in developing tropical locations. With the ease at which saliva is collected, the stability of these viral pathogens in saliva opens the possibility of remote sampling, and the use of at-home collection kits for diagnostics, as well as surveillance.

The surveillance of ARIs and respiratory viruses should be considered essential in this changing vaccine landscape. In addition to the regular updates to IAV and assessing those required for seasonal SARS-CoV-2 vaccines, several RSV vaccines are in the pipeline, with Arexvy and Abrysvo, already being rolled out in the U.S. for individuals over 60 years or pregnant individuals, and nirsevimab now approved for those under 24 months of age. With this, the population immune landscape is also changing. As such, surveillance efforts should not only be observing the outcome incidence among vaccine recipients, but also continuously assessing the impact of releasing these vaccines on the general population. Moreover, as SARS-CoV-2 demonstrated, virus genetic diversity can alter the effectiveness of control measures (Meredith et al., 2020). In the case of RSV for example, temporal changes in the circulating RSV strains may impact effectiveness if strains with different properties emerge in the post-licensure period. As such, whole virus genomes can be sequenced from virus-positive saliva samples collected through sustainable surveillance efforts to monitor and inform vaccination strategies.

This approach also has potential for the surveillance of pneumococcus. In the first study season, pneumococcal prevalence was lower than what is typically reported from culture-based carriage detection (Wyllie et al., 2023). While pneumococcal detection in the current study may have been enhanced due to the presence of respiratory symptoms (which can lead to a higher density of pneumococcus, thereby improving detection (Moore et al., 2012)), these preliminary findings would suggest this approach could be explored further as a low-resource means of testing for pneumococcus in the community, especially when combined with virus testing to better understand their interactions.

Our study was subject to several important limitations. First, the recommended saliva volume for collection was set at 0.5 mL, which has been proven more than sufficient for testing purposes. Unfortunately, a considerable proportion of the remaining specimens or remnant samples did not possess an adequate volume for conducting stability analyses from which multiple aliquots were required. Consequently, the reduced number of samples available for analysis in these early stages limited the statistical power of our findings. Regardless, the data generated are promising. Second, it has been difficult to obtain the necessary number of paired nasopharyngeal and saliva samples for formal clinical validation of the assay, as during seasonal epidemics, clinics can become overwhelmed and due to increasing testing aversion to swabs it can be difficult to recruit individuals to provide nasopharyngeal swabs for study purposes. Third, we observed significant differences between traditional extraction methods and extraction-free processing for the detection of hMPV in saliva. While testing extraction-free saliva lysates appeared comparable to testing nucleic acid extracted from nasal swabs, as compared to testing saliva following RNA extraction, further studies are required for improving the sensitivity of RNA-extraction-free detection of hMPV. However, it was interesting to observe an increase in hMPV-specific signal over the course of the stability study; disruption to the integrity of the sample over time improved the extraction-free detection of hMPV, the mechanism behind which should be further investigated. Finally, IAB was not detected during the study period. This was not surprising, as it was reported that only 0.5% of the influenza cases reported in the US at this time were identified as IAB (Merced-Morales et al., 2022). However, this meant that we were unable to assess the stability of IBV detection in raw, unsupplemented saliva.

Saliva, when collected and tested properly, has proven to be a reliable alternative sample type for SARS-CoV-2 detection (Tan et al., 2021). More importantly, we have demonstrated its potential for low-resource diagnostics and screening efforts, using an open-source RNA-extraction-free approach and the flexibility of this approach in outbreak response (Thomas et al., 2023). We further highlight the adaptability of this approach in the current study, demonstrating its potential for targeting other key respiratory pathogens, beyond SARS-CoV-2, namely, IAV, RSV and hMPV. While additional studies testing paired nasopharyngeal and saliva samples will help to further validate this approach for low-cost surveillance of respiratory viruses in the community and/or low-resource settings, this method holds promise for sustainable surveillance of respiratory pathogens in the community.

## Data Availability

All data produced in the present study are available upon reasonable request to the authors.

## DECLARATIONS

### Author’s contributions

ALW conceived the study. OAM and ALW managed the study. BT managed sample collection. OAM, TYL, KF, DY-C, JW and ALW were responsible for and performed the assays. OMA and ALW performed the analyses and interpreted the data. OMA, TYL, KF and ALW drafted the manuscript. All authors amended and commented on the final manuscript.

## Acknowledgements

We gratefully acknowledge labs for their efforts in supporting sample collection and the participants for their time and commitment to the study.

## REFERENCES

Allicock, O. M., Petrone, M. E., Yolda-Carr, D., Breban, M., Walsh, H., Watkins, A. E., Rothman, J. E., Farhadian, S. F., Grubaugh, N. D., & Wyllie, A. L. (2022). Evaluation of saliva self-collection devices for SARS-CoV-2 diagnostics. BMC Infect. Dis., 22(1), 284. 10.1186/s12879-022-07285-7

Bonroy, C., Vankeerberghen, A., Boel, A., & De Beenhouwer, H. (2007). Use of a multiplex real-time PCR to study the incidence of human metapneumovirus and human respiratory syncytial virus infections during two winter seasons in a Belgian paediatric hospital. Clin. Microbiol. Infect., 13(5), 504–509. 10.1111/j.1469-0691.2007.01682.x

Brandt, C. D., Kim, H. W., Arrobio, J. O., Jeffries, B. C., Wood, S. C., Chanock, R. M., & Parrott, R. H. (1973). Epidemiology of respiratory syncytial virus infection in Washington, DC: III. Composite analysis of eleven consecutive yearly epidemics. Am. J. Epidemiol., 98(5), 355–364.

Brueggemann, A. B., van Rensburg, M. J. J., Shaw, D., McCarthy, N. D., Jolley, K. A., Maiden, M. C., van Der Linden, M. P., Amin-Chowdhury, Z., Bennett, D. E., & Borrow, R. (2021). Changes in the incidence of invasive disease due to Streptococcus pneumoniae, Haemophilus influenzae, and Neisseria meningitidis during the COVID-19 pandemic in 26 countries and territories in the Invasive Respiratory Infection Surveillance Initiative: A prospective analysis of surveillance data. Lancet Digit. Health, 3(6), e360–e370.

CDC. (2020). CDC’s Influenza SARS-CoV-2 Multiplex Assay. https://www.cdc.gov/coronavirus/2019-ncov/lab/multiplex.html

CDC. (2022). US influenza surveillance: Purpose and methods. US Influenza Surveillance: Purpose and Methods. https://www.cdc.gov/flu/weekly/overview.htm

CDC. (2023). Current VISs. https://www.cdc.gov/vaccines/hcp/vis/current-vis.html

Cooney, M. K., Fox, J. P., & Hall, C. E. (1975). The Seattle virus watch: VI. Observations of infections with and illness due to parainfluenza, mumps and respiratory syncytial viruses and Mycoplasma pneumoniae. Am. J. Epidemiol., 101(6), 532–551.

Danino, D., Ben-Shimol, S., Van der Beek, B. A., Givon-Lavi, N., Avni, Y. S., Greenberg, D., Weinberger, D. M., & Dagan, R. (2022). Decline in pneumococcal disease in young children during the coronavirus disease 2019 (COVID-19) pandemic in Israel associated with suppression of seasonal respiratory viruses, despite persistent pneumococcal carriage: A prospective cohort study. Clin. Infect. Dis., 75(1), e1154–e1164.

Edwards, K. M., Zhu, Y., Griffin, M. R., Weinberg, G. A., Hall, C. B., Szilagyi, P. G., Staat, M. A., Iwane, M., Prill, M. M., & Williams, J. V. (2013). Burden of human metapneumovirus infection in young children. N. Engl. J. Med., 368(7), 633–643.

Fry, A. M., Chittaganpitch, M., Baggett, H. C., Peret, T. C. T., Dare, R. K., Sawatwong, P., Thamthitiwat, S., Areerat, P., Sanasuttipun, W., Fischer, J., Maloney, S. A., Erdman, D. D., & Olsen, S. J. (2010). The burden of hospitalized lower respiratory tract infection due to respiratory syncytial virus in rural Thailand. PLoS One, 5(11), e15098. 10.1371/journal.pone.0015098

Ginocchio, C. C., & McAdam, A. J. (2011). Current best practices for respiratory virus testing. J. Clin. Microbiol., 49(9_Supplement), S44–S48.

Hamid, S., Winn, A., Parikh, R., Jones, J. M., McMorrow, M., Prill, M. M., Silk, B. J., Scobie, H. M., & Hall, A. J. (2023). Seasonality of Respiratory Syncytial Virus—United States, 2017–2023. MMWR Morb Mortal Wkly Rep, 72(14), 355.

Hills, T., Kearns, N., Kearns, C., & Beasley, R. (2020). Influenza control during the COVID-19 pandemic. The Lancet, 396(10263), 1633–1634.

ISTA. (2007). ISTA 7D 2007 shipping standard. https://ista.org/docs/7Doverview.pdf

Kuitunen, I., Artama, M., Haapanen, M., & Renko, M. (2022). Respiratory virus circulation in children after relaxation of COVID-19 restrictions in fall 2021-A nationwide register study in Finland. J. Med. Virol., 94(9), 4528–4532. 10.1002/jmv.27857

Laxton, C. S., Peno, C., Hahn, A. M., Allicock, O. M., Perniciaro, S., & Wyllie, A. L. (2023). The potential of saliva as an accessible and sensitive sample type for the detection of respiratory pathogens and host immunity. Lancet Microbe. 10.1016/S2666-5247(23)00135-0

Li, Y., Reeves, R. M., Wang, X., Bassat, Q., Brooks, W. A., Cohen, C., Moore, D. P., Nunes, M., Rath, B., & Campbell, H. (2019). Global patterns in monthly activity of influenza virus, respiratory syncytial virus, parainfluenza virus, and metapneumovirus: A systematic analysis. Lancet Glob. Health, 7(8), e1031–e1045.

Li, Y., Wang, X., Cong, B., Deng, S., Feikin, D. R., & Nair, H. (2022). Understanding the potential drivers for respiratory syncytial virus rebound during the coronavirus disease 2019 pandemic. J. Infect. Dis., 225(6), 957–964.

Marom, T., Alvarez-Fernandez, P. E., Jennings, K., Patel, J. A., McCormick, D. P., & Chonmaitree, T. (2014). Acute bacterial sinusitis complicating viral upper respiratory tract infection in young children. Pediatr. Infect. Dis. J., 33(8), 803.

McLaughlin, J. M., Khan, F., Schmitt, H.-J., Agosti, Y., Jodar, L., Simões, E. A., & Swerdlow, D. L. (2022). Respiratory syncytial virus–associated hospitalization rates among US infants: A systematic review and meta-analysis. J. Infect. Dis., 225(6), 1100–1111.

Merced-Morales, A., Daly, P., Abd Elal, A. I., Ajayi, N., Annan, E., Budd, A., Barnes, J., Colon, A., Cummings, C. N., & Iuliano, A. D. (2022). Influenza activity and composition of the 2022–23 influenza vaccine—United States, 2021–22 season. MMWR Morb Mortal Wkly Rep, 71(29), 913.

Meredith, L. W., Hamilton, W. L., Warne, B., Houldcroft, C. J., Hosmillo, M., Jahun, A. S., Curran, M. D., Parmar, S., Caller, L. G., & Caddy, S. L. (2020). Rapid implementation of SARS-CoV-2 sequencing to investigate cases of health-care associated COVID-19: A prospective genomic surveillance study. Lancet Infect Dis., 20(11), 1263–1271.

Miller, E. K., Lu, X., Erdman, D. D., Poehling, K. A., Zhu, Y., Griffin, M. R., Hartert, T. V., Anderson, L. J., Weinberg, G. A., & Hall, C. B. (2007). Rhinovirus-associated hospitalizations in young children. J. Infect. Dis., 195(6), 773–781.

Monto, A. S. (2002). Epidemiology of viral respiratory infections. Am. J. Med., 112(6), 4–12.

Moore, D. P., Dagan, R., & Madhi, S. A. (2012). Respiratory viral and pneumococcal coinfection of the respiratory tract: Implications of pneumococcal vaccination. Expert Rev. Respir. Med., 6(4), 451–465.

Moriyama, M., Hugentobler, W. J., & Iwasaki, A. (2020). Seasonality of Respiratory Viral Infections. Annu Rev Virol, 7(1), 83–101. 10.1146/annurev-virology-012420-022445

Ott, I. M., Strine, M. S., Watkins, A. E., Boot, M., Kalinich, C. C., Harden, C. A., Vogels, C. B. F., Casanovas-Massana, A., Moore, A. J., Muenker, M. C., Nakahata, M., Tokuyama, M., Nelson, A., Fournier, J., Bermejo, S., Campbell, M., Datta, R., Dela Cruz, C. S., Farhadian, S. F., … the Yale IMPACT Research team3. (2021). Stability of SARS-CoV-2 RNA in Nonsupplemented Saliva. Emerg. Infect. Dis., 27(4), 1146–1150. 10.3201/eid2704.204199

Ramirez, J., Carrico, R., Wilde, A., Junkins, A., Furmanek, S., Chandler, T., Schulz, P., Hubler, R., Peyrani, P., Liu, Q., Trivedi, S., Uppal, S., Kalina, W. V., Falsey, A. R., Walsh, E. E., Yacisin, K., Jodar, L., Gessner, B. D., & Begier, E. (2023). Diagnosis of Respiratory Syncytial Virus in Adults Substantially Increases When Adding Sputum, Saliva, and Serology Testing to Nasopharyngeal Swab RT–PCR. Infect Dis Ther, 12(6), 1593–1603. 10.1007/s40121-023-00805-1

Tan, S. H., Allicock, O., Armstrong-Hough, M., & Wyllie, A. L. (2021). Saliva as a gold-standard sample for SARS-CoV-2 detection. Lancet Respir Med, 9(6), 562–564.

Tan, S. H., Allicock, O., Katamba, A., Carrington, C., Wyllie, A., & Armstrong-Hough, M. (2022). Saliva-based methods for SARS-CoV-2 testing in low- and middle-income countries. Bull. World Health Organ., 100(12), 808–814. 10.2471/BLT.22.288526

Thomas, R. J., Yolda-Carr, D., Fajardo, K., Allicock, O. M., Steel, S. A., Zepeda, T., Brownlee, M., Saladi, S., Parkin, J., & Wyllie, A. L. (2023). Expansion of a low-cost, saliva-based PCR test for the detection of mpox virus. medRxiv, 2023–06.

Tobik, E. R., Kitfield-Vernon, L. B., Thomas, R. J., Steel, S. A., Tan, S. H., Allicock, O. M., Choate, B. L., Akbarzada, S., & Wyllie, A. L. (2022). Saliva as a sample type for SARS-CoV-2 detection: Implementation successes and opportunities around the globe. Expert Rev. Mol. Diagn., 22(5), 519–535. 10.1080/14737159.2022.2094250

Trzciński, K., Bogaert, D., Wyllie, A., Chu, M. L. J. N., Van Der Ende, A., Bruin, J. P., Van Den Dobbelsteen, G., Veenhoven, R. H., & Sanders, E. A. M. (2013). Superiority of Trans-Oral over Trans-Nasal Sampling in Detecting Streptococcus pneumoniae Colonization in Adults. PLoS ONE, 8(3), e60520. 10.1371/journal.pone.0060520

Vogels, C. B. F., Watkins, A. E., Harden, C. A., Brackney, D. E., Shafer, J., Wang, J., Caraballo, C., Kalinich, C. C., Ott, I. M., Fauver, J. R., Kudo, E., Lu, P., Venkataraman, A., Tokuyama, M., Moore, A. J., Muenker, M. C., Casanovas-Massana, A., Fournier, J., Bermejo, S., … Yang, Y. (2021). SalivaDirect: A simplified and flexible platform to enhance SARS-CoV-2 testing capacity. Med, 2(3), 263–280.e6. 10.1016/j.medj.2020.12.010

Vos, T., Lim, S. S., Abbafati, C., Abbas, K. M., Abbasi, M., Abbasifard, M., Abbasi-Kangevari, M., Abbastabar, H., Abd-Allah, F., Abdelalim, A., Abdollahi, M., Abdollahpour, I., Abolhassani, H., Aboyans, V., Abrams, E. M., Abreu, L. G., Abrigo, M. R. M., Abu-Raddad, L. J., Abushouk, A. I., … Murray, C. J. L. (2020). Global burden of 369 diseases and injuries in 204 countries and territories, 1990–2019: A systematic analysis for the Global Burden of Disease Study 2019. Lancet, 396(10258), 1204–1222. 10.1016/S0140-6736(20)30925-9

Wyllie, A. L., Fournier, J., Casanovas-Massana, A., Campbell, M., Tokuyama, M., Vijayakumar, P., Warren, J. L., Geng, B., Muenker, M. C., Moore, A. J., Vogels, C. B. F., Petrone, M. E., Ott, I. M., Lu, P., Venkataraman, A., Lu-Culligan, A., Klein, J., Earnest, R., Simonov, M., … Ko, A. I. (2020). Saliva or Nasopharyngeal Swab Specimens for Detection of SARS-CoV-2. N Engl J Med, 383(13), 1283–1286. 10.1056/NEJMc2016359

Wyllie, A. L., Mbodj, S., Thammavongsa, D. A., Hislop, M. S., Yolda-Carr, D., Waghela, P., Nakahata, M., Stahlfeld, A. E., Vega, N. J., York, A., Allicock, O. M., Wilkins, G., Ouyang, A., Siqueiros, L., Strong, Y., Anastasio, K., Alexander-Parrish, R., Arguedas, A., Gessner, B. D., & Weinberger, D. M. (2023). Persistence of Pneumococcal Carriage among Older Adults in the Community despite COVID-19 Mitigation Measures. Microbiol. Spectr., 11(3), e04879–22. 10.1128/spectrum.04879-22

Wyllie, A. L., Wijmenga-Monsuur, A. J., van Houten, M. A., Bosch, A. A. T. M., Groot, J. A., van Engelsdorp Gastelaars, J., Bruin, J. P., Bogaert, D., Rots, N. Y., Sanders, E. A. M., & Trzciński, K. (2016). Molecular surveillance of nasopharyngeal carriage of Streptococcus pneumoniae in children vaccinated with conjugated polysaccharide pneumococcal vaccines. Sci. Rep., 6, 23809. 10.1038/srep23809

Yuan, H., Yeung, A., & Yang, W. (2022). Interactions among common non-SARS-CoV-2 respiratory viruses and influence of the COVID-19 pandemic on their circulation in New York City. Influenza Other Respir Viruses, 16(4), 653–661.

